# Chronic fatigue, depression and anxiety symptoms in Long COVID are strongly predicted by neuroimmune and neuro-oxidative pathways which are caused by the inflammation during acute infection

**DOI:** 10.1101/2022.06.29.22277056

**Authors:** Hussein Kadhem Al-Hakeim, Haneen Tahseen Al-Rubaye, Abbas F. Almulla, Dhurgham Shihab Al-Hadrawi, Michael Maes

## Abstract

**Background:** Long-term coronavirus disease 2019 (Long COVID) is associated with physio-somatic (chronic fatigue syndrome and somatic symptoms) and affective (depression and anxiety) symptoms. The severity of the Long COVID physio-affective phenome is largely predicted by peak body temperature (BT) and lowered oxygen saturation (SpO2) during the acute infectious phase. This study aims to delineate whether the association of BT and SpO2 during the acute phase and the Long COVID physio-affective phenome is mediated by neurotoxicity (NT) resulting from activated immune-inflammatory and oxidative stress pathways.

**Methods:** We recruited 86 patients with Long COVID (3-4 months after the acute phase) and 39 healthy controls and assessed serum C-reactive protein (CRP), caspase-1, interleukin (IL)-1β, IL-18, IL-10, myeloperoxidase (MPO), advanced oxidation protein products (AOPP), total antioxidant capacity (TAC), and calcium (Ca), as well as peak BT and SpO2 during the acute phase.

**Results:** Cluster analysis revealed that a significant part (34.9%) of Long COVID patients (n=30) show a highly elevated NT index computed based on IL-1β, IL-18, Caspase-1, CRP, MPO and AOPP. Partial Least Squares analysis showed that 61.6% of the variance in the physio-affective phenome of Long COVID is explained by the NT index, lowered Ca, peak BT/SpO2 in the acute phase, and prior vaccinations with Astra-Zeneca or Pfizer. The most important predictors of the physio-affective phenome are Ca, CRP, IL-1β, AOPP and MPO.

**Conclusion:** The infectious-immune-inflammatory core of acute COVID-19 strongly predicts the development of physio-affective symptoms 3-4 months later, and these effects are partly mediated by neuro-immune and neuro-oxidative pathways.

## Introduction

While the infection rate with severe acute respiratory syndrome coronavirus 2 (SARS-CoV-2) fell globally (Moghadas, Vilches et al. 2021, Andrews, Stowe et al. 2022), a new concern arose among many post-infected individuals having Long COVID symptoms (Lerner, Robinson et al. 2021). Several studies reported a cluster of persistent symptoms extending beyond full recovery of coronavirus disease (COVID-19) (Deng, Zhou et al. 2021, Huang, Huang et al. 2021, Lopez-Leon, Wegman-Ostrosky et al. 2021). These symptoms either appear following up to 2 (infection-related), 3 (acute-post-COVID), or 6 (prolonged post-COVID) months or more (chronic post-COVID) after the acute infectious phase (Fernández-de-Las-Peñas, Palacios-Ceña et al. 2021). Many individuals (74-.87.4%) with Long COVID suffer from a variety of mental and physiosomatic symptoms after recovery from the acute phase (Carfì, Bernabei et al. 2020, Arnold, Hamilton et al. 2021), including chronic fatigue, affective symptoms (low mood and anxiety), cognitive dysfunctions and sleep disturbances, along with somatic manifestations such as autonomic symptoms, muscle pain, muscle tension, headache, a flu-like malaise, gastro-intestinal symptoms (GIS), shortening of breath, persistent cough, and chest pain (Cirulli, Schiabor Barrett et al. 2020, Garrigues, Janvier et al. 2020, Arnold, Hamilton et al. 2021, Bellan, Soddu et al. 2021, Cares-Marambio, Montenegro-Jiménez et al. 2021, Davis, Assaf et al. 2021, Deng, Zhou et al. 2021, Huang, Huang et al. 2021, Renaud-Charest, Lui et al. 2021, Sandler, Wyller et al. 2021, Shah, Hillman et al. 2021, Simani, Ramezani et al. 2021, van den Borst, Peters et al. 2021).

The immunopathogenesis of the acute infectious phase of COVID-19 comprises activation of the cytokine network with induction of interferons, and interleukin (IL)-1, IL-6, IL-12, IL-18, and tumor necrosis factor (TNF)-α signaling, activation of the nucleotide-binding domain, leucine-rich repeat and pyrin domain-containing protein 3 (NLRP3) inflammasome and as well as activation of key antiviral pathways, Toll-Like-Receptor cascades, NOD-like receptor signaling (Yalcinkaya, Liu et al. 2021, Zhao, Di et al. 2021, Maes, Tedesco Junior et al. 2022, Sagulkoo, Plaimas et al. 2022). The NLRP3 is a key component of the innate immune system and an intracellular sensor that is induced by pathogen-associated molecular patterns and damage-associated molecular patterns (Junqueira, Crespo et al. 2021, Vora, Lieberman et al. 2021). NLRP3 activation causes increased levels of caspase-1, IL-1β, and IL-18 and cell death or pyroptosis (Swanson, Deng et al. 2019, Rodrigues, de Sá et al. 2021). Increased peak body temperature (BT) and lowered oxygen saturation (SpO2) reflect the severity of the immune-inflammatory response during acute COVID and predict critical COVID-19 and increased mortality (Tharakan, Nomoto et al. 2020, Al-Hadrawi, Al-Rubaye et al. 2022, Maes 2022).

Recently, we introduced a new concept, namely “the physio-affective phenome of acute and Long COVID” to describe the physio-somatic (fatigue and somatic symptoms) and affective (depression and anxiety) symptoms in both illnesses [review (Al-Hadrawi, Al-Rubaye et al. 2022, Al-Hakeim, Al-Rubaye et al. 2022, Al-Jassas, Al-Hakeim et al. 2022)]. Thus, in acute and Long COVID, a validated latent construct could be derived from depressive, anxiety, chronic fatigue and multiple physio-somatic symptoms and, therefore, this factor score reflects the severity of the intertwined increases in physio-somatic and affective symptoms (Al-Hadrawi, Al-Rubaye et al. 2022, Al-Hakeim, Al-Rubaye et al. 2022, Al-Jassas, Al-Hakeim et al. 2022).

In acute COVID-19, this physio-affective phenome was largely explained by the cumulative effects of increased pro-inflammatory cytokines, including IL-6, soluble advanced glycation products (sRAGEs), changes in acute phase proteins, including C-reactive protein (CRP) and albumin, lowered calcium (Ca), pneumonia as indicated by chest computerized tomography scan abnormalities (CCTAs), and diminished SpO2 (Al-Jassas, Al-Hakeim et al. 2022). Moreover, elevated peak BT and lowered SpO2 during the acute phase of infection predict the physio-affective phenome of Long COVID (Al-Hadrawi, Al-Rubaye et al. 2022). In Long COVID, the severity of the physio-affective phenome was also predicted by increased oxidative toxicity as indicated by increased levels of malondialdehyde (MDA), protein carbonyls (PCs), myeloperoxidase (MPO), and nitric oxide (NO), and lowered antioxidant defenses as indicated by lowered zinc and glutathione peroxidase (Gpx) (Al-Hakeim, Al-Rubaye et al. 2022). Moreover, the oxidative toxicity / antioxidant ratio was significantly predicted by increased peak BT and lowered SpO2 (Al-Hakeim, Al-Rubaye et al. 2022). These results indicate that the immune-inflammatory response in the acute COVID-19 predicts, at least in part, the physio-affective symptoms of Long COVID and that these effects are partly mediated by increased neuro-oxidative toxicity (Al-Hakeim, Al-Rubaye et al. 2022, Al-Jassas, Al-Hakeim et al. 2022).

It is important to note that neuro-immune and nitro-oxidative toxicity play a key role in the pathophysiology of chronic fatigue syndrome (CFS), major depression (MDD), and generalized anxiety disorder (GAD) (Maes, Bonifacio et al. 2018, Maes and Carvalho 2018, Almulla and Maes 2022, Maes, Kubera et al. 2022). There is now also evidence that upregulation of the NLRP3 inflammasome plays a key role in MDD (Kaufmann, Costa et al. 2017, Zhou, Fernando et al. 2021), fatigue (Zhang, Du et al. 2016, Zhang, Ma et al. 2017), cognitive impairments (Kuwar, Rolfe et al. 2021), and anxiety (Aghaie, Moradifar et al. 2021, Smith, Trageser et al. 2021). Nevertheless, there are no data whether the NLRP3 inflammasome and lowered Ca are involved in the physio-affective phenome of Long COVID.

Hence, this study was performed to delineate a) the effects of the NLRP3 inflammasome (IL-1β, IL-18 and caspase 1), and inflammatory (CRP), oxidative stress (advanced oxidation protein products, AOPP and MPO) and total antioxidant capacity (TAC) and lowered Ca on the physio-affective phenome of Long COVID; and b) whether the neuro-immune and neuro-oxidative toxicity is predicted by lowered SpO2 and increased peak BT during the acute phase. The specific hypotheses are that the physio-affective phenome of Long COVID is predicted by lowered SpO2 and increased peak BT and that these effects are at least in part mediated by increased neurotoxicity (NT) due to increased caspase-1, IL-1β, and IL-18, CRP, MPO, AOPP and lowered TAC and Ca levels. The data are analyzed using the new precision nomothetic psychiatry approach (Maes 2022, Maes and Stoyanov 2022) which involves the construction of a new endophenotype class of Long COVID.

## Participants and Methods

### Participants

We used the World health organization (WHO) criteria to diagnose Long COVID patients (World Health Organization 2021). These criteria are a) daily life activities of the patients should be influenced by minimally two symptoms, namely; fatigue, impairment of memory or concentration, achy muscles, absence of smell or taste senses, affective symptoms, and cognitive impairment, b) patients should have a confirmed infection with COVID-19, c) the symptoms should persist beyond the acute phase or should became apparent 2-3 months later; and d) the symptoms should last for at least two months (World Health Organization 2021) and are present 3-4 months after recovery. Accordingly, 86 Long COVID patients participated in the current study, clustered into two groups: Long COVID with low (n=56) and high (n=30) NT (as defined below). Additionally, we recruited 39 controls. All participants have been recruited from September to the end of December 2021. The present study involved a combined methodology namely the impact of acute COVID-19 phase on Long COVID symptoms (retrospective study design) and a comparison of abnormalities in Long COVID patients versus healthy control (case-control design).

In the acute COVID-19 phase, specialized clinicians and virologists diagnosed patients with a positive COVID-19 infection based on: a) severe symptoms of infection, such as fever, cough, shortness of breath, and the loss of smell and taste senses, and b) positive test results of reverse transcription real-time polymerase chain reaction (rRT-PCR) findings test, and c) positive immunoglobulin M (IgM) against SARS-COV-2. All patients were quarantined and treated at several hospitals and specialized centers within Al-Najaf city; namely Al-Sader Medical City of Najaf, Al-Hakeem General Hospital, Al-Zahraa Teaching Hospital for Maternity and Pediatrics, Imam Sajjad Hospital, Hassan Halos Al-Hatmy Hospital for Transmitted Diseases, Middle Euphrates Center for Cancer, and Al-Najaf teaching hospital.

All participants with Long COVID were free of any signs of acute COVID-19, namely dry cough, sore throat, shortness of breath, fever, night sweats, or chills and before participating in the current study, patients and controls showed negative rRT-PCR test results. Around 33% of the participants in the control group had minor mental symptoms such as low mood, anxiety, and fatigue resulting from the quarantine period and lack of social activities, which may also affect patients with Long COVID. We excluded subjects with a previous major depressive episode, bipolar disorder, dysthymia, GAD, panic disorder, schizo-affective disorder, schizophrenia, psycho-organic syndrome, and substance use disorders (except tobacco use disorder, TUD), neurodegenerative and inflammatory diseases such as CFS (Morris and Maes 2013), Parkinson’s or Alzheimer’s disease, multiple sclerosis, stroke, or systemic (auto)immune diseases such as diabetes mellitus, psoriasis, rheumatoid arthritis, inflammatory bowel disease and scleroderma. We also excluded subjects with liver and renal disease, and pregnant and breastfeeding women.

The present study was designed and performed in line with Iraqi and international ethical and privacy laws, including the World Medical Association’s Declaration of Helsinki, The Belmont Report, the Council for International Organizations of Medical Sciences (CIOMS) Guideline, and the International Conference on Harmonization of Good Clinical Practice; our institutional review board adheres to the International Guidelines for Human Research Safety (ICH-GCP). We obtained written consent from all participants, parents, or any legally responsible person prior involvement in our study. The institutional ethics board and the Najaf Health Directorate-Training and Human Development Center approved our research according to their documents which numbered (8241/2021) and (18378/ 2021), respectively.

### Clinical measurements

Following 3-4 months after recovery from acute COVID-19, a semi-structured interview was conducted by a senior psychiatrist to obtain the socio-demographic and clinical characteristics of all participants. The psychiatrist assessed several symptom domains of chronic fatigue and fibromyalgia symptoms utilizing the Fibro-fatigue (FF) scale (Zachrisson, Regland et al. 2002), severity of depression using the Hamilton Depression Rating Scale (HAMD) (Hamilton, 1960) and the Beck Depression Inventory - II (BDI-II) (Hautzinger 2009), and severity of anxiety symptoms using the Hamilton Anxiety Rating Scale (HAMA) (Hamilton 1959). Moreover, we utilized those rating scale items to derive subdomains of the major symptoms. We made two subdomains of the HAMD; the first is pure depressive symptoms (pure HAMD) which is the sum of sad mood, feelings of guilt, suicidal thoughts, and loss of interest; and the second, physiosomatic HAMD (physiosom HAMD) which is the sum of somatic anxiety, gastrointestinal (GIS) anxiety, genitourinary anxiety, and hypochondriasis. Likewise, two subdomains of the HAMA were computed, namely pure anxiety symptoms (pure HAMA) as the sum of anxious mood, tension, fears, anxiety and anxious behavior during the interview, and physiosomatic HAMA symptoms (physiosom HAMA) as the sum of somatic sensory, cardiovascular, GIS, genitourinary, and autonomic symptoms. Furthermore, after omitting items reflecting cognitive and affective symptoms in the FF scale, a pure physiosomatic FF (pure FF) score was computed as the sum of muscular pain, muscle tension, fatigue, autonomous symptoms, GIS symptoms, headache and a flu-like malaise. We also computed the sum of all pure depressive BDI-II (pure BDI) symptoms thus excluding physiosomatic symptoms, including sadness, discouraged about the future, feeling a failure, dissatisfaction, feeling guilty, feeling punished, disappointed in self, critical of self, suicidal ideation, crying, loss of interest, difficulties with decisions, and work inhibition. The physio-affective phenome was defined in our previous studies (Al-Hadrawi, Al-Rubaye et al. 2022, Al-Jassas, Al-Hakeim et al. 2022) as the first factor extracted from pure FF and BDI, and pure and physiosom HAMA and HAMD scores. The DSM-5 criteria were used to diagnose TUD. Weight in kilograms was divided by height in meters squared to calculate the body mass index (BMI).

We used the patients’ records to obtain the lowest SpO2 and peak BT values, which were obtained during hospitalization for the acute infectious phase. The assessments were made by a well-trained paramedical person who employed an electronic oximeter manufactured by Shenzhen Jumper Medical Equipment Co. Ltd. and a sublingual digital thermometer provided with beep sound. By subtracting z transformed SpO2 values from z transformed peak BT values, we generated a new index involving both lowered SpO2 and increased peak BT (dubbed: TO2 index). Additionally, we recorded the different types of vaccines received by patients, namely AstraZeneca, Pfizer and Sinopharm.

### Assays

Following an overnight fasting at 7.30-9.00 a.m., five milliliters of venous blood were drawn utilizing disposable syringes and transferred directly to serum tubes. We avoided any hemolyzed, lipemic, and icteric blood samples. All tubes were centrifuged at 3000 rpm after 10 minutes incubation at room temperature. Then, we made three aliquots of serum that were stored at -70 LJ in Eppendorf tubes until thawed for biochemical analyses. We employed ELISA kits provided by Nanjing Pars Biochem Co.,Ltd. (Nanjing, China) to assess serum levels of IL-1β, IL-18, IL-10, caspase-1, MPO, TAC and AOPP (albumin ratio). Total serum Ca was measured spectrophotometrically using ready-to-use kits obtained from Agappe Diagnostics Ltd., Cham, Switzerland. We used a z unit-based composite score to assess a new NT index by computing z transformation of IL-1β (z IL-1β) + z IL-18 + z caspase-1 + z MPO + z AOPP + z CRP.

### Statistical analysis

In the present study, IBM SPSS software version 28 was used to carry out all statistics. We conducted analysis of variance (ANOVA) to delineate the differences in continuous variables among study groups and analysis of contingency tables to examine associations between nominal variables. Pearson product-moment correlation coefficients were used to analyze the relationship between two scale variables. Multivariate and univariate general linear models (GLM) were used to examine the association between clinical and biomarker data and the diagnostic categories, while allowing or controlling for age, TUD, sex, BMI, and education. Estimated marginal mean (SE) values were computed and multiple group mean differences were assessed using Fisher’s protected (the omnibus test is significant) Least Significant Difference (LSD). The ability of biomarkers and clinical variables to predict the physio-affective symptoms was determined by multiple regression analysis. We utilized an automated stepwise approach with 0.05 p-value and 0.06 to include and omit, respectively. For each one of the explanatory variables, we computed the standardized beta-coefficients, t statistics, and the exact p-value along with F statistics and total variance explained (R^2^) for the model. Furthermore, we used the variance inflation factor and tolerance to examine multicollinearity. The heteroskedasticity was checked by employing the White and modified Breusch-Pagan tests. We used cluster analysis (two-step) and followed the precision nomothetic approach (Maes 2022) to construct endophenotype classes of patients with Long COVID based on a combination of SpO2, peak BT and the neurotoxic biomarkers in a z unit-based composite score (z IL-1β + z IL-18 + z caspase-1 + z MPO + z AOPP + z CRP), dubbed NT index. The cluster solution was considered adequate when the silhouette measure of cohesion and separation was > 0.5. Canonical correlation analysis was employed to investigate the correlations between two sets of variables, namely physio-affective symptoms following 3-4 months after acute COVID infection as the dependent variables and biomarkers as explanatory variables. The variance explained by the canonical variables of both sets was computed as well as the variance in the canonical dependent variable set explained by the independent canonical variable set. We accepted the canonical components when the explained variance of both sets was > 0.50 and when all canonical loadings were > 0.5.

Partial Least Squares (PLS) analysis was used to study the causative relationships between SARS-CoV-2 infection, peak BT and lowest SpO2 during the acute phase of disease and the physio-affective phenome of Long COVID, whereby the effects of the input variables are partly mediated by NT and other biomarkers. All input variables were inputted as single indicators, and the output variable was a latent vector extracted from the values of pure and physiosom HAMA and HAMD and pure FF and BDI (the physio-affective phenome). Complete PLS analysis was conducted only when the outer and inner models met the following prespecified quality criteria: a) all loadings on the extracted latent vector are > 0.6 at p < 0.001, b) the output latent vector shows high construct and convergence validity as indicated by rho A >0.8, Cronbach’s alpha >0.7, composite reliability >0.7, and average variance extracted (AVE) > 0.5, c) blindfolding demonstrates that the construct’s cross-validated redundancy is sufficient, d) Confirmatory Tetrad Analysis (CTA) demonstrates that the latent vector extracted from the rating scale scores is not mis-specified as a reflective model, e) the model’s prediction performance is satisfactory as measured by PLS Predict, and f) the model fit is <0.08 in terms of standardized root squared residual (SRMR) values. If all model quality data conform with the prespecified criteria, we conduct a complete PLS-SEM pathway analysis with 5000 bootstrap samples and calculate the path coefficients (with p-values) as well as specific and total indirect (mediated) effects and total effects.

## Results

### Socio-demographic data

We employed a two-steps cluster analysis to classify the patients with Long COVID into two groups employing peak BT, SpO2 and the NT index (IL-1β + IL-18 + Caspase-1+ MPO + AOPP+ CRP), with the aim to developed a new biomarker-derived endophenotype class within the Long COVID patient group. Entered in the cluster analysis were the diagnosis of Long COVID as nominal variable and peak BT, lowered SpO2 and the NT composite score as the continuous variables. According to the silhouette measure of cohesion and separation of 0.53, the quality of the clusters was adequate. Three clusters were derived, namely the healthy control sample (n= 39), and patients with high peak BT, lowered SpO2 and an increased NT index (dubbed as high TO-NT, T for temperature, O for SpO2 and NT for neurotoxicity) (n=30) and patients (n= 56) with less pronounced changes in these biomarkers (dubbed low TO-NT). Therefore, Long COVID patients were classified based on the combination of two acute COVID-19 phase markers and blood biomarkers 3-4 months after clinical recovery.

Socio-demographic and clinical data of the three groups are presented in **Table 1**. SpO2, peak BT, the TO2 index and NT index were significantly different between the high and low TO-NT clusters. No significant changes were detected between these groups in age, sex, BMI, marital state, smoking, residency, vaccination state, and education.

**Table 1.** Socio-demographic data, body temperature (BT), and oxygen saturation (SpO2) in healthy controls (HC) and Long COVID patients, classified according to peak body temperature (BT), oxygen saturation (SpO2) and the neurotoxicity (NT) index as Long COVID with high (high TO-NT) versus low (low TO-NT) levels of these biomarkers.

| Variables | HC (n=39)<sup>A</sup> | Low TO-NT (n=56)<sup>B</sup> | High TO-NT (n=30)<sup>C</sup> | F/X<sup>2</sup> | df | P |
| --- | --- | --- | --- | --- | --- | --- |
| Age (Years) | 28.3 (7.6) | 27.6 (5.4) | 29.8 (7.3) | 1.15 | 2/122 | 0.320 |
| Sex (M/F) | 42 / 15 | 40 / 16 | 22 / 8 | 1.42 | 2 | 0.512 |
| Marital state (Ma/S) | 22 / 17 | 31 / 25 | 18 / 12 | 0.18 | 2 | 0.946 |
| Smoking (Y/N) | 13 / 26 | 16 / 40 | 11 / 19 | 0.64 | 2 | 0.732 |
| Residency (U/R) | 31 / 8 | 46 / 10 | 25 / 5 | 0.19 | 2 | 0.914 |
| Vaccination (A/Pf/S) | 9 / 21 / 9 | 14 / 31 / 11 | 6 / 17 / 7 | 0.42 | 4 | 0.979 |
| BMI (Kg/m<sup>2</sup>) | 25.60 (3.97) | 25.71 (3.74) | 26.96 (5.75) | 0.99 | 2/122 | 0.372 |
| Education (Years) | 15.0 (1.3) | 15.7 (1.8) | 15.5 (1.7) | 2.73 | 2/122 | 0.069 |
| Peak BT (°C) | 36.86 (0.25)<sup>B,C</sup> | 38.20 (0.64)<sup>A,C</sup> | 39.31 (0.83)<sup>A,B</sup> | 138.38 | 2/122 | <0.001 |
| Lowest SpO<sub>2</sub> (%) | 95.08 (1.52)<sup>B,C</sup> | 92.41 (2.74)<sup>A,C</sup> | 88.27 (4.62)<sup>A,B</sup> | 42.81 | 2/122 | <0.001 |
| TO<sub>2</sub> index (z scores) | -1.01 (0.240)<sup>B,C</sup> | 0.054 (0.52)<sup>A,C</sup> | 1.21 (0.86)<sup>A,B</sup> | 131.43 | 2/122 | <0.001 |
| Composite NT (z score) | -0.595 (0.85)<sup>B,C</sup> | -0.143 (0.80)<sup>A,C</sup> | 1.04 (0.66)<sup>A,B</sup> | 37.91 | 2/122 | <0.001 |

Results are shown as mean (SD): F: results of analysis of variance; X<sup>2</sup>: analysis of contingency tables. <sup>A,B,C</sup>: Results of pairwise comparisons among group means. TO-NT; (T)Temperature, (O)Oxygen saturation,(NT)Neurotoxic index, M: Male, F: Female, Ma: Married, S: Single, U: Urban, R: Rural, BMI: Body Mass Index, A: AstraZeneca, Pf: Pfizer, S: Sinopharm, Y: Yes, N: No, TO<sub>2</sub> index: computed as z BT – z SpO<sub>2</sub>. NT: neurotoxicity computed as a z unit-based composite score

### Differences in psychiatric rating scales between study groups

**Table 2** shows the measurements of the rating scales, namely the total and subdomains scores. There were significant differences among the three study groups in total and pure HAMD, HAMA, BDI, FF and physiosom HAMD and HAMA. Table 2 shows that the scores of all scales and subscales, except pure HAMA, increased from controls to the low TO-NT group to the high TO-NT group. The pure HAMA score was not significantly different between the two patients groups but was higher in the latter than in controls.

**Table 2.** Clinical rating scales scores in healthy controls (HC) and Long COVID patients classified according to peak body temperature (BT), oxygen saturation (SpO2) and the neurotoxicity (NT) index as Long COVID with high versus low levels of these biomarkers (dubbed TO-NT).

| Variables | HC (n=39) <sup>A</sup> | Low TO-NT (n=56) <sup>B</sup> | High TO-NT (n=30) <sup>C</sup> | F/X<sup>2</sup><br>df=2/119 | p |
| --- | --- | --- | --- | --- | --- |
| Total HAMD | 5.5 (0.67) <sup>B,C</sup> | 15.7 (0.6) <sup>A,C</sup> | 19.1 (0.8) <sup>A,B</sup> | 105.21 | <0.001 |
| Total BDI | 8.6 (1.0) <sup>B,C</sup> | 23.5 (0.9) <sup>A,C</sup> | 27.0 (1.2) <sup>A,B</sup> | 85.35 | <0.001 |
| Total HAMA | 7.8 (1.1) <sup>B,C</sup> | 15.4 (0.9) <sup>A,C</sup> | 19.5 (1.3) <sup>A,B</sup> | 26.23 | <0.001 |
| Total FF | 10.9 (1.8) <sup>B,C</sup> | 24.7 (1.5) <sup>A,C</sup> | 35.4 (2.0) <sup>A,B</sup> | 43.05 | <0.001 |
| Pure FF (z score) | -0.877 (0.122) <sup>B,C</sup> | 0.146 (0.101) <sup>A,C</sup> | 0.867 (0.138) <sup>A,B</sup> | 46.06 | <0.001 |
| Pure HAMD (z score) | -0.964 (0.118) <sup>B,C</sup> | 0.296 (0.098) <sup>A,C</sup> | 0.702 (0.133) <sup>A,B</sup> | 51.04 | <0.001 |
| Pure BDI (z score) | -1.030 (0.113) <sup>B,C</sup> | 0.348 (0.094) <sup>A,C</sup> | 0.700 (0.128) <sup>A,B</sup> | 62.79 | <0.001 |
| Pure HAMA (z score) | -0.606 (0.139) <sup>B,C</sup> | 0.190 (0.115) <sup>A</sup> | 0.432 (0.157) <sup>A</sup> | 14.37 | <0.001 |
| Physiosom HAMA (z score) | -0.495 (0.145) <sup>B,C</sup> | 0.029 (0.120) <sup>A,C</sup> | 0.588 (0.164) <sup>A,B</sup> | 12.21 | <0.001 |
| Physiosom HAMD (z score) | -1.031 (0.118) <sup>B,C</sup> | 0.347 (0.098) <sup>A,C</sup> | 0.702 (0.133) <sup>A,B</sup> | 57.93 | <0.001 |

All results of univariate GLM analysis; data are expressed as estimated marginal mean (SE) values obtained by GLM analysis after covarying for age, sex, education, and smoking. <sup>A,B,C</sup>: Results of pairwise comparisons among group means
FF: Fibro fatigue scale, HAMA: Hamilton Anxiety Rating Scale, HAMD: Hamilton Depression Rating Scale, BDI: Beck Depression Inventory-II, Physiosom: physio-somatic

### Differences in biomarkers between study groups

The measurements of the biomarkers in both classes of patients with Long COVID versus healthy controls are displayed in **Table 3.** CRP and AOPP were significantly different between the three study groups. Caspase 1 was significantly higher in the high TO-NT class than in the two other groups. IL-1β was significantly higher in the high TO-NT group than in controls and IL-10 higher in both Long COVID groups than in controls. Total Ca was lower in patients than in controls.

**Table 3.**
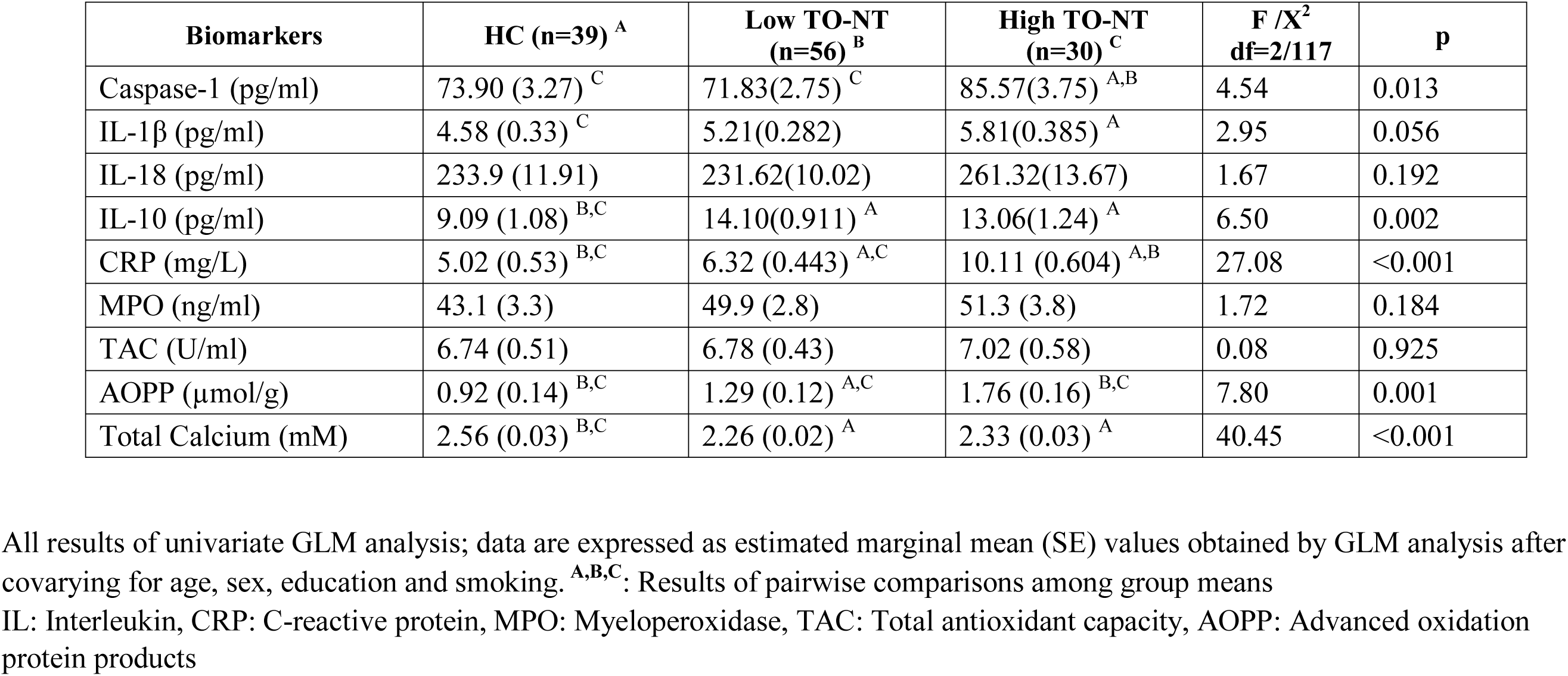
**B**iomarkers in healthy controls (HC) and Long COVID patients classified according to peak body temperature (BT), oxygen saturation (SpO2) and the neurotoxicity (NT) index as Long COVID with high (high TO-NT) versus low (low TO-NT) levels of these biomarkers.

| **Biomarkers** | **HC (n=39) <sup>A</sup>** | **Low TO-NT (n=56) <sup>B</sup>** | **High TO-NT (n=30) <sup>C</sup>** | **F /X<sup>2</sup>**<br>**df=2/117** | **p** |
| --- | --- | --- | --- | --- | --- |
| Caspase-1 (pg/ml) | 73.90 (3.27) <sup>C</sup> | 71.83(2.75) <sup>C</sup> | 85.57(3.75) <sup>A,B</sup> | 4.54 | 0.013 |
| IL-1β (pg/ml) | 4.58 (0.33) <sup>C</sup> | 5.21(0.282) | 5.81(0.385) <sup>A</sup> | 2.95 | 0.056 |
| IL-18 (pg/ml) | 233.9 (11.91) | 231.62(10.02) | 261.32(13.67) | 1.67 | 0.192 |
| IL-10 (pg/ml) | 9.09 (1.08) <sup>B,C</sup> | 14.10(0.911) <sup>A</sup> | 13.06(1.24) <sup>A</sup> | 6.50 | 0.002 |
| CRP (mg/L) | 5.02 (0.53) <sup>B,C</sup> | 6.32 (0.443) <sup>A,C</sup> | 10.11 (0.604) <sup>A,B</sup> | 27.08 | <0.001 |
| MPO (ng/ml) | 43.1 (3.3) | 49.9 (2.8) | 51.3 (3.8) | 1.72 | 0.184 |
| TAC (U/ml) | 6.74 (0.51) | 6.78 (0.43) | 7.02 (0.58) | 0.08 | 0.925 |
| AOPP (μmol/g) | 0.92 (0.14) <sup>B,C</sup> | 1.29 (0.12) <sup>A,C</sup> | 1.76 (0.16) <sup>B,C</sup> | 7.80 | 0.001 |
| Total Calcium (mM) | 2.56 (0.03) <sup>B,C</sup> | 2.26 (0.02) <sup>A</sup> | 2.33 (0.03) <sup>A</sup> | 40.45 | <0.001 |

All results of univariate GLM analysis; data are expressed as estimated marginal mean (SE) values obtained by GLM analysis after covarying for age, sex, education and smoking. <sup>A,B,C</sup>: Results of pairwise comparisons among group means IL: Interleukin, CRP: C-reactive protein, MPO: Myeloperoxidase, TAC: Total antioxidant capacity, AOPP: Advanced oxidation protein products

### Prediction of the clinical rating scales

**Table 4** shows multiple regression analyses with total and subdomains rating scale scores as dependent variables and biomarkers as explanatory variables. In regression #1 and #2 we introduced the first PC extracted from pure FF, HAMD, HAMA, BDI and physiosom HAMD and HAMA scores (dubbed PC Physio-Affective phenome score, reflecting overall severity) as dependent variable. We found that 46.0% of the variance in this PC (regression #1) was explained by total Ca, the NT index and BMI. **Figures 1** and **2** show the partial regression of PC physio-affective score on total Ca and the NT index, respectively. After introducing peak BT and SpO2 in regression #2, we found that a large part of the variance (52.4%) in PC physio-affective phenome was explained by peak BT and NT (both positively associated). **Figure 3** shows the partial regression of the phenome score on peak BT.

**Figure 1.**
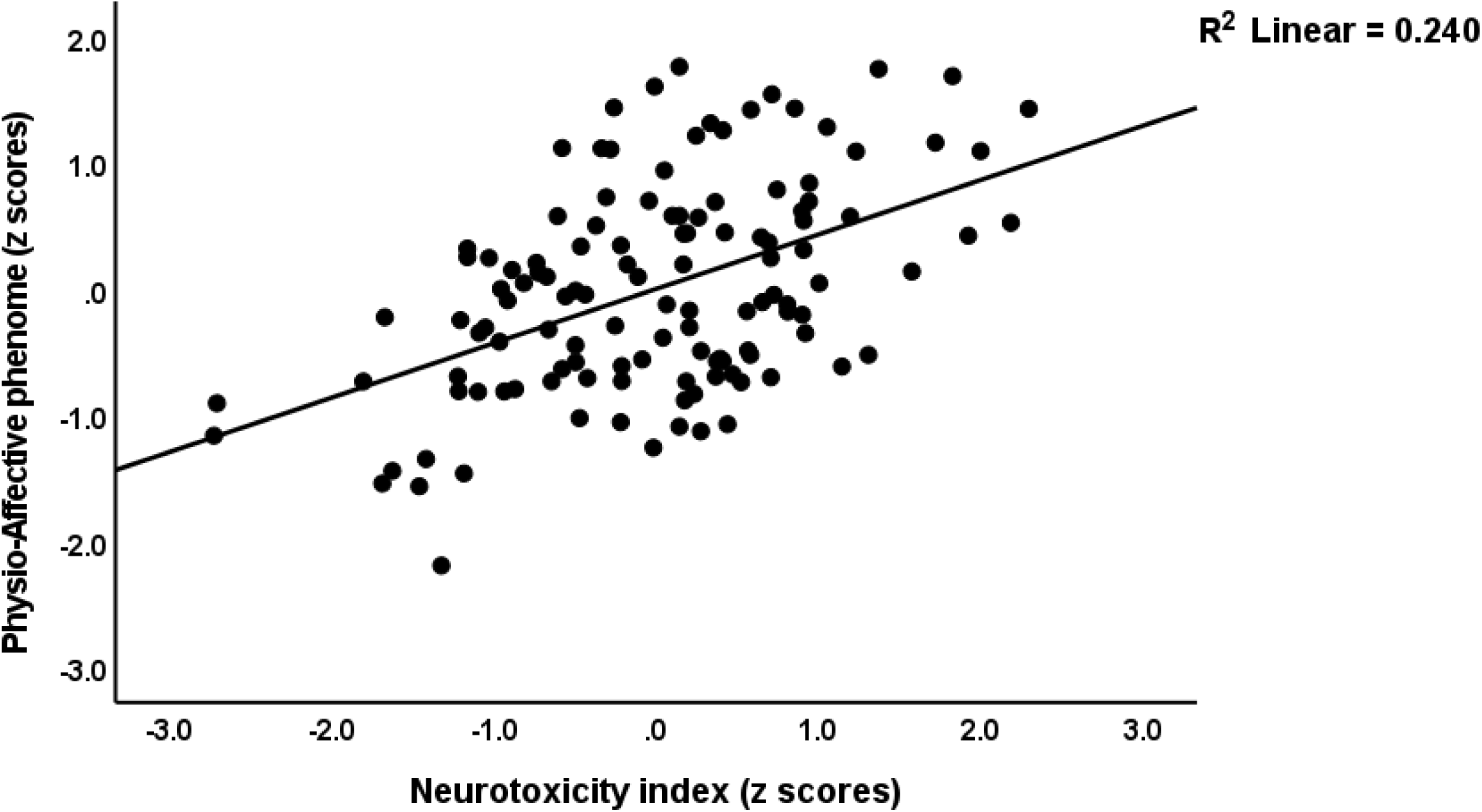
Partial regression of the physio-affective phenome score in Long COVID patients and healthy controls on the neurotoxicity index (IL-1β + IL-18 + Caspase-1+ MPO + AOPP+ CRP).

**Figure 2.**
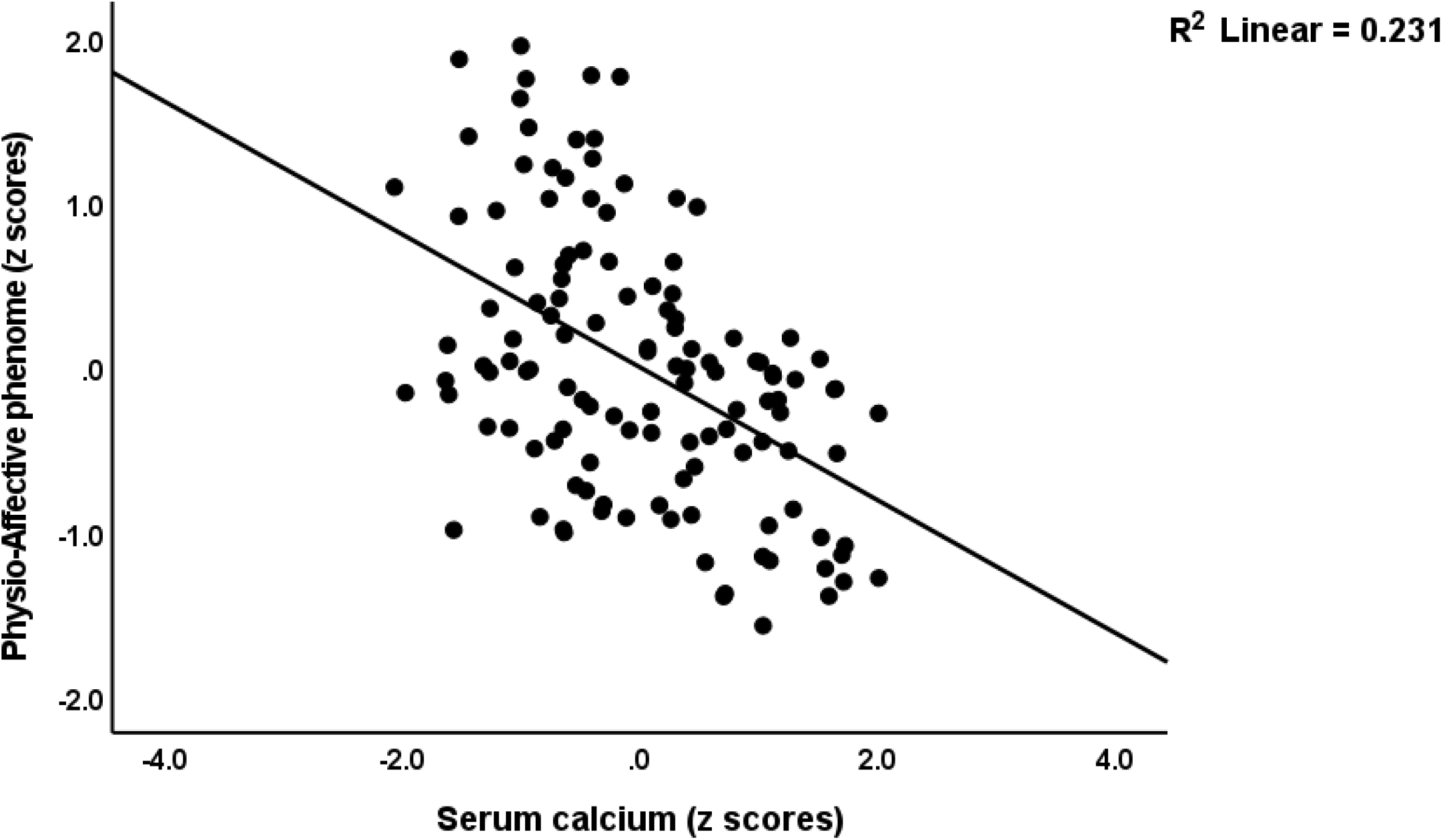
Partial regression of the physio-affective phenome score in Long COVID patients and healthy controls on serum total calcium.

**Figure 3.**
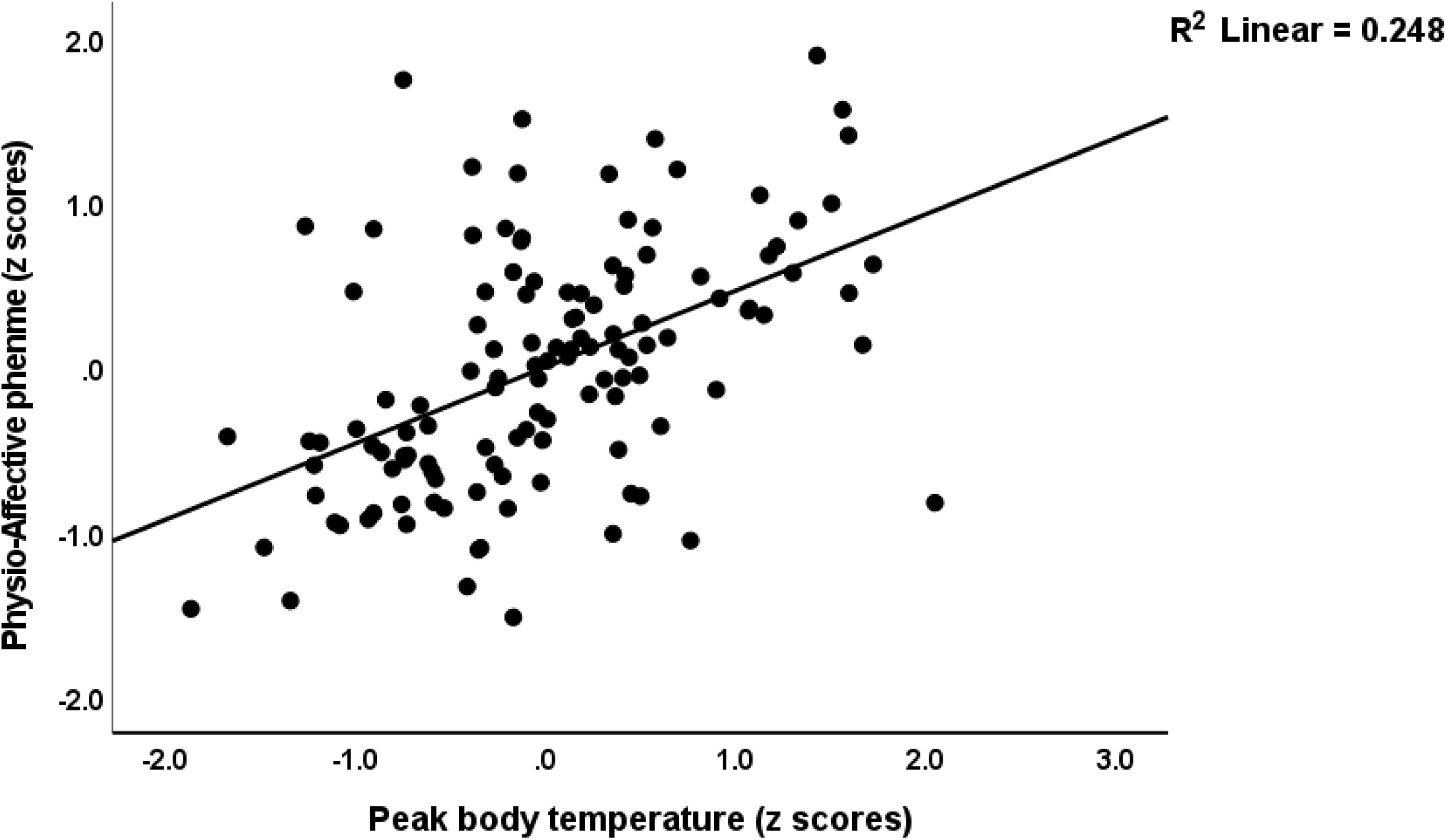
Partial regression of the physio-affective phenome score in Long COVID patients and healthy controls on peak body temperature.

**Table 4:** Results of multiple regression analyses with different physio-somatic and affective rating scale scores as dependent variables and peak body temperature (BT) and oxygen saturation (SpO2), neurotoxicity (NT) and biomarkers as explanatory variables.

| **Dependent Variables** | **Explanatory Variables** | **B** | **t** | **p** | **Model R<sup>2</sup>** | **F** | **df** | **p** |
| --- | --- | --- | --- | --- | --- | --- | --- | --- |
| **#1 PC Physio-somatic phenome** | **Model** |  |  |  | 0.460 | 34.06 | 3/120 | <0.001 |
|  | Total calcium | -0.414 | -6.00 | <0.001 |  |  |  |  |
|  | NT | 0.420 | 6.16 | <0.001 |  |  |  |  |
|  | BMI | 0.164 | 2.42 | 0.017 |  |  |  |  |
| **#2 PC Physio-somatic phenome** | **Model** |  |  |  | 0.574 | 53.88 | 3/120 | <0.001 |
|  | Peak BT | 0.472 | 6.29 | <0.001 |  |  |  |  |
|  | Calcium | -0.253 | -3.76 | <0.001 |  |  |  |  |
|  | NT | 0.229 | 3.38 | <0.001 |  |  |  |  |
| **#3 Pure FF** | **Model** |  |  |  | 0.371 | 11.61 | 6/118 | <0.001 |
|  | Total Calcium | -0.346 | -4.56 | <0.001 |  |  |  |  |
|  | CRP | 0.242 | 3.09 | 0.002 |  |  |  |  |
|  | Education | 0.192 | 2.59 | 0.011 |  |  |  |  |
|  | AOPP | 0.214 | 2.74 | 0.007 |  |  |  |  |
|  | Vaccination Astra | 0.166 | 2.23 | 0.027 |  |  |  |  |
|  | BMI | 0.149 | 1.99 | 0.048 |  |  |  |  |
| **#4 Pure HAMD** | **Model** |  |  |  | 0.413 | 13.86 | 6/118 | <0.001 |
|  | Total Calcium | -0.381 | -5.25 | <0.001 |  |  |  |  |
|  | CRP | 0.224 | 2.97 | 0.004 |  |  |  |  |
|  | Education | 0.244 | 3.37 | 0.001 |  |  |  |  |
|  | Vaccination Astra | 0.186 | 2.58 | 0.011 |  |  |  |  |
|  | AOPP | 0.201 | 2.67 | 0.009 |  |  |  |  |
|  | MPO | 0.155 | 2.11 | 0.037 |  |  |  |  |
| **#5 Pure BDI** | **Model** |  |  |  | 0.372 | 14.12 | 5/119 | <0.001 |
|  | Total Calcium | -0.368 | -4.87 | <0.001 |  |  |  |  |
|  | AOPP | 0.247 | 3.17 | 0.002 |  |  |  |  |
|  | Education | 0.197 | 2.61 | 0.010 |  |  |  |  |
|  | CRP | 0.218 | 2.80 | 0.006 |  |  |  |  |
| | Interleukin-1β | 0.174 | 2.22 | 0.028 | | | | |
| **#6 Pure HAMA** | **Model** |  |  |  | 0.183 | 9.06 | 3/121 | <0.001 |
|  | Total Calcium | -0.296 | -3.52 | 0.001 |  |  |  |  |
|  | CRP | 0.204 | 2.42 | 0.017 |  |  |  |  |
|  | MPO | 0.170 | 2.06 | 0.041 |  |  |  |  |
| **#7 Physiosom**<br>**HAMD** | **Model** |  |  |  | 0.374 | 17.88 | 4/120 | <0.001 |
|  | Total Calcium | -0.432 | -5.84 | <0.001 |  |  |  |  |
|  | CRP | 0.301 | 4.07 | <0.001 |  |  |  |  |
|  | Interleukin-18 | 0.181 | 2.49 | 0.014 |  |  |  |  |
|  | Vaccination Astra+Pfizer | 0.159 | 2.19 | 0.030 |  |  |  |  |
| **#8 Physiosom**<br>**HAMA** | **Model** |  |  |  | 0.217 | 11.17 | 3/121 | <0.001 |
|  | Total Calcium | -0.301 | -3.70 | <0.001 |  |  |  |  |
|  | MPO | 0.231 | 2.85 | 0.005 |  |  |  |  |
|  | BMI | 0.207 | 2.53 | 0.013 |  |  |  |  |

FF: Fibro-Fatigue scale, HAMA: Hamilton Anxiety Rating Scale, HAMD: Hamilton Depression Rating Scale, BDI: Beck Depression Inventory, Physiosom: Physio-somatic, BMI: Body Mass Index, CRP: C-reactive protein, AOPP: Advanced oxidation protein products, MPO: Myeloperoxidase

In all regression analyses shown in this table (regressions 3-7), NT was with Ca the most significant predictor. Therefore, we show the solutions without the NT index to allow for the effects of the separate NT biomarkers. We found that 37.1% of the variance in pure FF scores (regression #3) could be explained by total Ca (inversely), CRP, education, AOPP, BMI, and vaccination with AstraZeneca (all positively associated). The results of regression #4 revealed that total Ca (inversely), CRP, education, AOPP, MPO, and vaccination with Astra Zeneca (all positively) predicted 41.3% of the variance in pure HAMD scores. We found that (regression #5) total Ca (inversely associated), CRP, education, AOPP, and IL-1β (all positively associated) explained 37.2% of the variance in the pure BDI score. Regression #6 shows that a significant part of the variance (18.3%) in pure HAMA could be predicted by total Ca (inversed associated), CRP, and MPO (both positively associated). The results of regression #7 indicate that in Long COVID patients, 37.4% of the variance in physiosom HAMD was explained by total Ca (inversely), vaccination with Pfizer or Astra Zeneca, CRP, and IL-18 (all positively associated). Regression #8 shows that total Ca (inversely associated), MPO and BMI (both positively associated) explained 21.7 % of the variance in physiosom HAMA scores.

### Results of canonical correlations

We employed canonical correlation analyses to detect the association between a set of explanatory variables, namely the NT index, TO2 index, and total Ca, and various symptom domains of Long COVID as dependent variables (**Table 5)**. The canonical components extracted from NT composite, TO2, and total Ca strongly correlated with physio-affective symptoms and could explain 35.5% of the variance.

**Table 5.** Results of canonical correlation analyses examining the associations between two sets of variables, namely set 1: the physio-somatic and affective rating scale scores as dependent variables, and set 2: neurotoxicity (NT), a composite based on peak body temperature and SpO2 (TO2 index) and serum calcium as explanatory variables

| Sets | Variables | Canonical Loadings |
| --- | --- | --- |
| **Set 1**<br>**Dependent** | Pure FF | 0.845 |
|  | Pure HAMD | 0.815 |
|  | Physiosom HAMD | 0.784 |
|  | Pure HAMA | 0.585 |
|  | Physiosom HAMA | 0.629 |
|  | Pure BDI | 0.825 |
| **Set 2**<br>**Explanatory** | NT Composite | 0.670 |
|  | TO<sub>2</sub> index | 0.875 |
|  | Total Calcium | 0.674 |
| **Statistics** | F (df) | 8.675 (18/325) |
|  | P | <0.001 |
|  | Correlation | 0.799 |
|  | Set 1 / Set 2 | 0.355 |
|  | Set 2 by self | 0.556 |
|  | Set 1 by self | 0.569 |

FF: Fibro-Fatigue scale, HAMA: Hamilton Anxiety Rating Scale, HAMD: Hamilton Depression Rating Scale, BDI: Beck Depression Inventory, Physiosom: Physio-somatic, NT: a z unit-based composite score reflecting neurotoxicity; TO2: computed as z peak BT – SpO2

### Results of PLS analysis

**Figure 4** depicts a first PLS model that evaluated whether the effects of SpO2 and peak BT (introduced as a single indicator, namely the TO2 index) on the physio-affective phenome of Long COVID (entered as a latent vector taken from the 6 rating scale subdomains) are mediated via NT and Ca (IL-10 and TAC were not significant). Since the multiple regression analysis also showed effects of vaccination, we entered vaccination with Sinopharm as an additional explanatory variable. With an SRMR of 0.045, the model quality was satisfactory, and we observed appropriate construct reliability validity values for the physio-affective phenome with AVE = 0.613; rho A = 0.920; composite reliability = 0.904; and Cronbach alpha = 0.873; whilst all loadings of the 6 indicators of the physio-affective phenome were > 0.7. CTA showed that the latter vector was not mis-specified as a reflective model, and blindfolding indicated an acceptable construct cross-validated redundancy of 0.364. PLSPredict showed that the construct indicators had positive Q2 predict values, indicating that the prediction error was smaller than the naivest benchmark. Complete PLS path analysis showed that 61.6% of the variance in the physio-affective phenome was explained by the regression on NT, Ca, TO2 index, and vaccination and that the TO2 index explained 16.2% and 17.1% of the variances in NT and Ca, respectively. SARS-CoV-2 infection explained 47.0% of the variance in the TO2 index. While TO2 has significant direct effects on the phenome, it has also significant specific indirect effects either mediated via NT (t=4.10, p<0.001) or Ca (t=4.13, p<0.001). Infection results in a highly significant total indirect effect on the Long COVID phenome (t=8.92, p<0.001).

**Figure 4.**
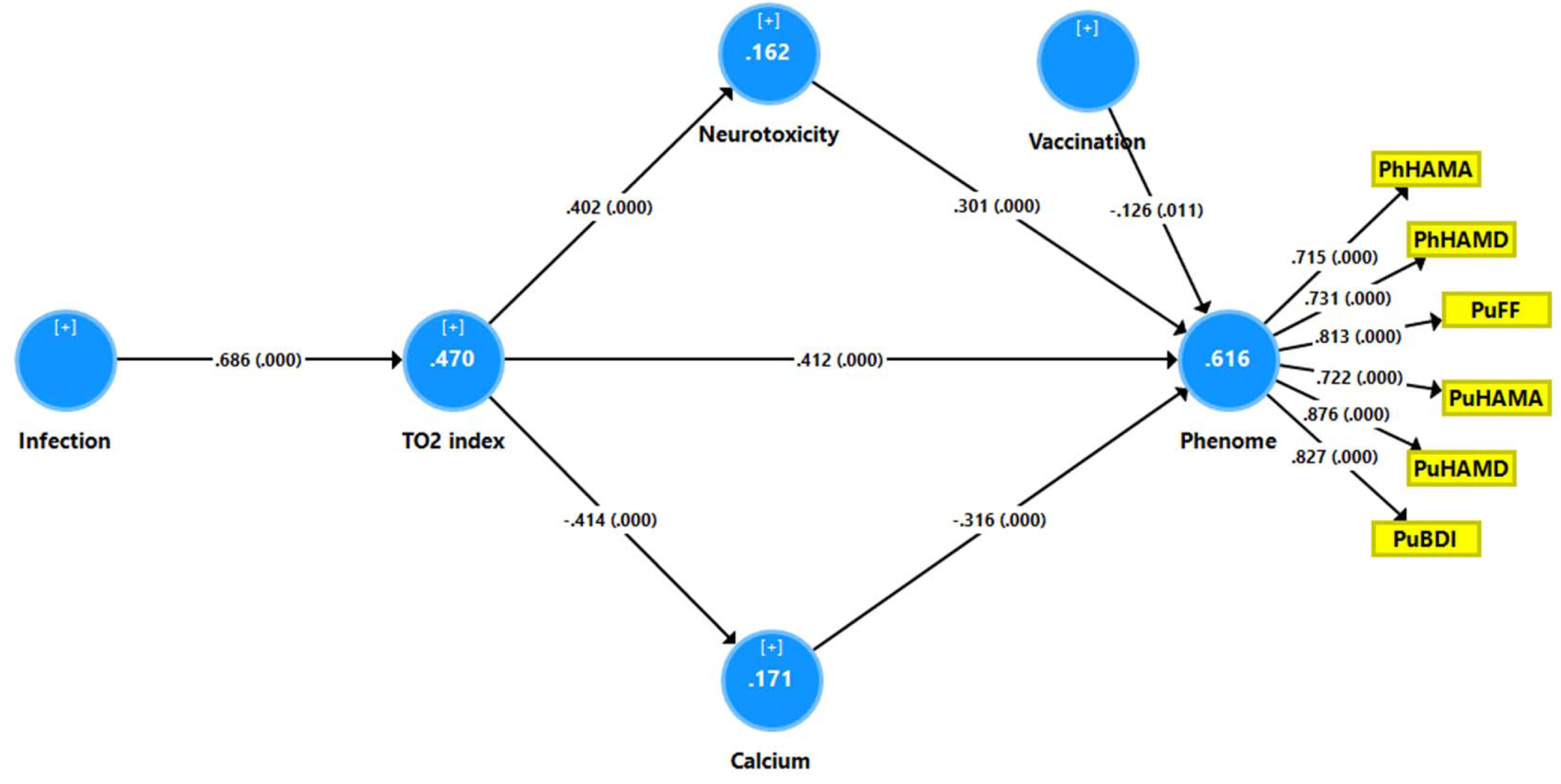
Results of Partial Least Squares (PLS)-SEM analysis with the physio-affective phenome score as the output variable. The latter is entered as a latent vector extracted from 6 symptom domains. The phenome latent vector is predicted by calcium, neurotoxicity index, the TO2 index and vaccination (Astra-Zeneca and Pfizer are coded as 0, Sinopharm as 1). SARS-CoV-2 infection is the primary input variable. As such, neurotoxicity and calcium partially mediate of the effects of TO2 and SARS-CoV-2 infection on the physio-affective phenome. Neurotoxicity: z unit-based composite score computed based on six neurotoxic immune and oxidative products. TO2: a z unit-based composite score computed as z BT – z SpO2. Vaccination: indicates that Astra-Zeneca and Pfizer vaccinations are positively associated with the phenome. PhHAMA: Physio-somatic symptoms of the Hamilton Anxiety Rating Scale; PhHAMD: physio-somatic symptoms of the Hamilton Depression Rating Scale, PuFF: Pure fatigue and physio-somatic symptoms of the Fibrofatigue scale, PuHAMA: Pure anxiety symptoms of the Hamilton Anxiety Rating Scale, PuHAMD: Pure depression symptoms of the Hamilton Depression Rating Scale, PuBDI: Pure depression scores from the Beck Depression Inventory. Vaccination: indicates that Astra-Zeneca and Pfizer vaccinations are positively associated with the phenome.

In order to examine the effects of SpO2 and peak BT on the separate biomarkers of Long COVID and to examine which biomarkers are the most important in predicting the phenome, we conducted a second PLS path analysis (see **Figure 5**). With an SRMR of 0.040, the model quality was adequate, and the construct reliability validity of the latent construct was adequate (not shown as similar to that explained in Figure 4). We found that 46.8% of the variance was explained by the regression on Ca, CRP, IL-1β, AOPP, MPO and vaccination. SpO2 has significant effects on AOPP and MPO, whilst peak BT affected MPO, CRP and Ca.

**Figure 5.**
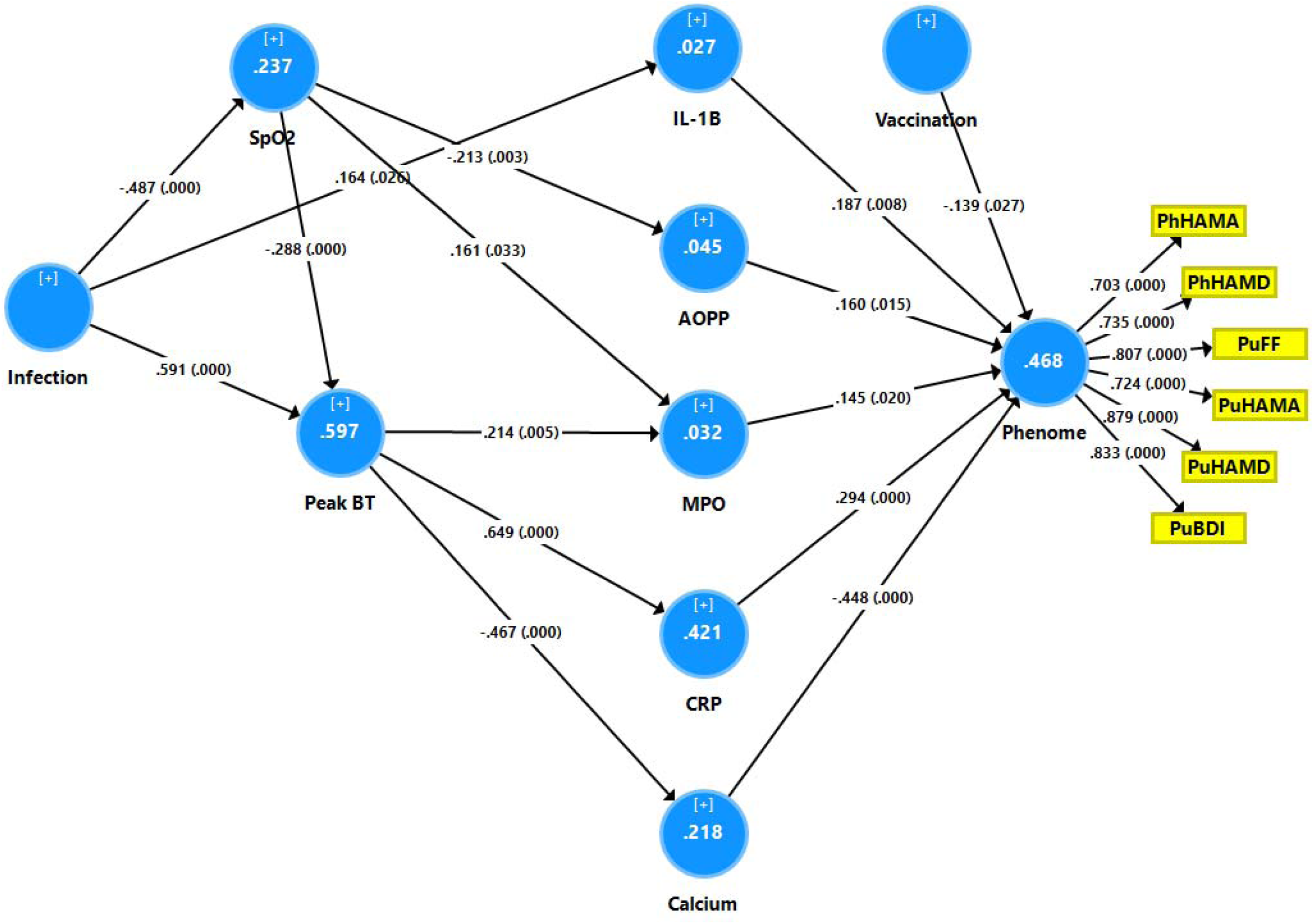
Results of Partial Least Squares (PLS)-SEM analysis with the physio-affective phenome score as the output variable. The latter is entered as a latent vector extracted from 6 symptom domains. The phenome latent vector is predicted by calcium, all neurotoxicity biomarkers, namely interleukin (IL)-1β, C-reactive protein (CRP), myeloperoxidase (MPO), and advanced oxidation protein products, peak body temperature (BT), oxygen saturation (SpO2) and vaccination (Astra-Zeneca and Pfizer are coded as 0, Sinopharm as 1). SARS-CoV-2 infection is the primary input variable. As such, the neurotoxicity biomarkers and calcium mediate the effects of BT, SpO2 and infection on the phenome. PhHAMA: Physio-somatic symptoms of the Hamilton Anxiety Rating Scale; PhHAMD: physio-somatic symptoms of the Hamilton Depression Rating Scale, PuFF: Pure fatigue and physio-somatic symptoms of the Fibrofatigue scale, PuHAMA: Pure anxiety symptoms of the Hamilton Anxiety Rating Scale, PuHAMD: Pure depression symptoms of the Hamilton Depression Rating Scale, PuBDI: Pure depression scores from the Beck Depression Inventory. Vaccination: indicates that Astra-Zeneca and Pfizer vaccinations are positively associated with the phenome.

## Discussion

### The physio-affective phenome of Long COVID

The first major finding of this study is that we were able to extract one replicable latent vector from the physio-somatic and affective rating scale scores. These findings confirm the results of another study conducted on an independent study sample of Iraqi COVID-19 patients and controls (Al-Hakeim, Al-Rubaye et al. 2022). Moreover, both the latter and the current study reported that the physio-affective core of Long COVID was strongly predicted by the combined effects of increased peak BT and lowered SpO2 during the acute phase of disease. As explained previously (Al-Jassas, Al-Hakeim et al. 2022), increased peak BT and lowered SpO2 reflect the severity of the infection-immune-inflammatory core of acute COVID-19. These results indicate that the physio-affective core during acute and Long COVID is largely the consequence of infection-immune-inflammatory pathways. The results confirm that physio-somatic symptoms including chronic fatigue, physio-somatic symptoms including pain, GIS, malaise, and autonomic symptoms and affective symptoms share common immune-inflammatory pathways as reviewed in the introduction.

In accordance with our previous study (Al-Hakeim, Al-Rubaye et al. 2022), we found that AstraZeneca (viral vector, genetically engineered virus) and Pfizer (mRNA) vaccination, but not administration of the Sinopharm vaccine (inactivated virus), may exacerbate the physio-affective phenome of Long COVID. Those SARS-CoV-2 vaccinations were reported in a few studies to be associated with Long COVID–like symptoms, including sadness, anxiety, exhaustion, and immune disoders, including increased synthesis of spike protein, T cell activation, deficits in type 1 interferon signaling, and autoimmune responses (Couzin-Frankel and Vogel 2022, Seneff, Nigh et al. 2022).

### Increased NT due to NLRP3 activation predicts the physio-affective phenome

The second major finding is that the present study constructed a new endophenotype class based on increased NT during Long-COVID, and lowered SpO2 and increased peak BT during the acute phase of illness (this cluster was dubbed TO-NT Long COVID). The latter cluster of patients is characterized by increased indicants of NLRP3 inflammasome activation with increased IL-1β and caspase 1, mild inflammation with increased CRP, increased MPO and AOPP and lower total Ca levels. It should be underscored that the non-TO-NT cluster of patients also showed increased NT, although significantly less than the TO-NT cluster. Although IL-10, a negative immunoregulatory cytokine, was significantly increased in Long COVID, IL-10 did not predict the phenome after considering the other biomarkers. Most importantly, patients belonging to the TO-NT cluster showed highly significant increases in physio-affective scores, indicating strong associations between biomarkers of acute and Long COVID and physio-affective symptoms. Moreover, the NT index and the key components of this index, namely increased IL-1β, CRP, MPO and AOPP and lowered Ca, predict a large part of the variance in the physio-affective phenome. As such, this study has reified the physio-affective phenome of Long COVID into a more concrete NT-driven concept.

It is interesting to note that NLRP3 genetic variants (namely NLPR3 rs10157379 T > C and NLPR3 rs10754558 C > G variants) are associated with fatigue, myalgia, hyperalgesia, and malaise in the acute infectious phase (Maes, Tedesco Junior et al. 2022). Abnormal NLRP3 activation during acute infection may lead to pathological tissue injury (Zhao, Di et al. 2021) and may underpin the exaggerated immune response since it contributes to the cytokine storm in acute COVID-19 (Tisoncik Jennifer, Korth Marcus et al. 2012, Lin, Xu et al. 2019). Several studies have reported that caspase-1, IL-1β, and IL-18 are associated with depression, anxiety, and fatigue, indicating the implication of the NLRP3 inflammasome in the pathophysiology of these diseases (Hardcastle, Brenu et al. 2015, Swartz, Prather et al. 2017, Towers, Oelschlager et al. 2017, Li, Zheng et al. 2018, VanElzakker, Brumfield et al. 2019). NLRP3 can activate the caspase-1 enzyme, which in turn triggers IL-1β and IL-18 pro-inflammatory cytokines to induce pyroptosis (cell death in response to pro-inflammatory signals) and plays a key role in neuroinflammation (Song, Pei et al. 2017).

IL-1β is necessary to start and maintain the immune-inflammatory reactions in the central nervous system (CNS) and may impact the integrity of blood brain barrier (BBB), leading to leakage of the peripheral immune cells into the CNS (Cannon 2000, Alvarez, Dodelet-Devillers et al. 2011). Moreover, IL-1β mediates microglia and astrocyte activation resulting in infiltration of T cells to the CNS, thus augmenting the pro-inflammatory state by producing IL-6 and TNF-α along with neurotoxic metabolites enhancing excitotoxicity and neuronal damage (Ferrari, Depino et al. 2004, Almulla and Maes 2022). IL-18 is one of the mediators of cell-mediated immunity, which triggers T helper (Th)-1 and B cells to generate adhesion molecules, pro-inflammatory cytokines, and chemokines (Nakahira, Ahn et al. 2002, Bossù, Ciaramella et al. 2010). In addition, IL-18 may increase microglial expression of caspase-1, matrix metalloproteinases, and the formation of pro-inflammatory cytokines (Felderhoff-Mueser, Schmidt et al. 2005) and may cause neuronal damage through elevations in Fas ligands in glial cells (Song, Pei et al. 2017). High IL-1β and IL-18 levels were detected in patients with CNS infection, brain injuries, Alzheimer’s disease, and multiple sclerosis (Licastro, Pedrini et al. 2000, de Jong, Huizinga et al. 2002, Huang, Huang et al. 2004). All in all, activation of the NLRP3 has neurotoxic effects either by promoting the synthesis of other detrimental metabolites or by damaging neurons directly.

### Increased NT due to increased CRP predicts the physio-affective core

In accordance with our previous study (Al-Hakeim, Al-Rubaye et al. 2022), we found that mild elevations in CRP contributed to the physio-affective phenome of Long COVID. Previous studies showed elevated CRP in COVID-19 patients following 2-3 months of full recovery (Gameil, Marzouk et al. 2021, Mandal, Barnett et al. 2021). In the liver, CRP production is triggered by IL-6 (Kuta and Baum 1986) and elevated CRP has toxic effects on endothelial cells and raises the permeability of BBB, thus enhancing the development of neurodegenerative and cerebrovascular diseases (Song, Kim et al. 2009, Windgassen, Funtowicz et al. 2011, Hsuchou, Kastin et al. 2012, Ge, Xu et al. 2013, Belin, Devic et al. 2020). For example, elevated CRP is associated with the impaired functional outcome and increased mortality due to stroke (Di Napoli, Papa et al. 2001, Masotti, Ceccarelli et al. 2005, Elkind, Tai et al. 2006, Montaner, Fernandez-Cadenas et al. 2006). Furthermore, studies showed increased CRP levels in depression (Howren, Lamkin et al. 2009, Roomruangwong, Kanchanatawan et al. 2017), suicidal behavior (Vasupanrajit, Jirakran et al. 2022), GAD (Costello, Gould et al. 2019), CFS (Strawbridge, Sartor et al. 2019) and cognitive impairment (Arce Rentería, Gillett et al. 2020).

### Increased NT due to oxidative stress predicts the physio-affective phenome

The third major finding of the current study is that the physio-affective phenome is associated with increased oxidative stress as indicated by increased MPO and AOPP, although in contrast to our prior hypothesis, TAC was not decreased in Long COVID. The present finding extend our previous results, revealing high nitro-oxidative stress and lowered antioxidant defenses (lowered zinc and Gpx levels) in Long COVID patients (Al-Hakeim, Al-Rubaye et al. 2022).

Neutrophils produce MPO enzyme as part of the innate immune response which may induce the generation of reactive chlorine species (RCS), such as hydrochlorous acid (Mathy-Hartert, Bourgeois et al. 1998), which may lead to chlorinative stress with the formation of AOPP (Maes, Landucci Bonifacio et al. 2019, da Cruz Nizer, Inkovskiy et al. 2020). Not only neutrophils but also microglia and pyramidal neurons of the hippocampus express a substantial amount of MPO enzyme, which is associated with disease conditions such as Alzheimer’s disease and multiple sclerosis (Green, Mendez et al. 2004, Gray, Thomas et al. 2008). Increased MPO and AOPP are reported in depression, anxiety, and cognitive impairment (Vaccarino, Brennan et al. 2008, Liang, Li et al. 2013, Talarowska, Szemraj et al. 2015, Shrivastava, Chelluboina et al. 2021).

Moreover, we observed that lowered SpO2 in the acute phase predicted increased AOPP and MPO in Long COVID, and that increased peak BT in the acute phase predicted increased MPO and CRP and lowered Ca in Long COVID. Likewise, our previous results (Al-Hakeim, Al-Rubaye et al. 2022) revealed that lowered SpO2 in the acute phase predicted lowered Gpx and increased NO production in Long COVID patients, and that elevated BT during acute COVID-19 predicted increased CRP and lowered antioxidant defenses including zinc in Long COVID. This indicates that the effects of the infection-immune-inflammatory core of acute COVID-19 on the physio-affective phenome of Long COVID is partly mediated by the cumulative effects of neuro-immune, neuro-oxidative and neuro-nitrosative pathways.

### Lowered total Ca levels predict the physio-affective phenome

The fourth major finding of the current study is that total Ca levels were lower in Long COVID than in controls and that lowered Ca significantly predicted the severity of the physio-affective phenome. In the acute COVID phase, there is a reduction in Ca which is strongly associated with the physio-affective core (Al-Jassas, Al-Hakeim et al. 2022). Previous studies reported that MDD and depressive symptoms are accompanied by significant reductions in ionized and total Ca levels (Bowden, Huang et al. 1988, Paul 2001, Jung, Ock et al. 2010, Islam, Islam et al. 2018, Al-Dujaili, Al-Hakeim et al. 2019). Ca levels are necessary to maintain normal mood and cognition by affecting neuronal signaling pathways and protecting neuroplasticity processes (Toescu and Verkhratsky 2007, Hurst 2010, Grützner, Listunova et al. 2018). Moreover, the lowered Ca levels in acute COVID-19 are part of the infection-immune-inflammatory core (Al-Jassas, Al-Hakeim et al. 2022), which also appears to determine lower Ca levels during Long COVID. Thus, it is safe to hypothesize that in patients with Long COVID, abnormal Ca which accompanies the immune-inflammatory response during the acute phase of illness, contributes mechanistically to the physio-affective phenome of both acute and Long COVID.

### Limitations

The findings of the current study should be interpreted regarding its limitations. First, the results would be more interesting if we had assessed biomarkers of chlorinative stress, namely chlorotyrosine and dichlorotyrosine, and activation of the tryptophan catabolite (TRYCAT) pathway which may increase TRYCATs and lower the availability of tryptophan, the precursor of 5-HT (Maes, Leonard et al. 2011, Almulla, Thipakorn et al. 2022). Moreover, our COVID-19 studies have been conducted on Iraqi patients and, therefore, deserve replication in other countries.

## Conclusions

High peak BT and lowered SpO2 during the acute phase of COVID-19 are associated with the development of the physio-affective phenome of Long COVID disease, and these effects are partially explained by increased NT through activation of the NLRP3 inflammasome, a mild inflammatory response, increased chrorinative stress and lowered total Ca levels. The infectious-immune-inflammatory core of acute COVID-19, and the development of NLRP3, a mild chronic inflammatory response, increased nitro-oxidative stress, lowered antioxidant defenses and total Ca levels are new drug targets to treat the physio-affective symptoms of Long COVID.

## Data Availability

The dataset generated during and/or analyzed during the current study will be available from the corresponding author (M.M.) upon reasonable request and once the dataset has been fully exploited by the authors.

## Acknowledgments

The authors would like to express their appreciation to the individuals who worked diligently to compile the data at the Al-Sader Medical City of Najaf, Al-Hakeem General Hospital, Al-Zahraa Teaching Hospital for Maternity and Pediatrics, Imam Sajjad Hospital, Hassan Halos Al-Hatmy Hospital for Transmitted Diseases, Middle Euphrates Center Cancer, and Al-Najaf Center for Cardiac Surgery and Trans Catheter.

## Ethical approval and consent to participate

The approval of the study was obtained from the institutional ethics board of the University of Kufa (8241/2021) and the Najaf Health Directorate-Training and Human Development Center (Document No.18378/ 2021). The present study was performed under Iraqi and foreign ethics and privacy rules besides the following guidelines; World Medical Association Declaration of Helsinki, The Belmont Report, CIOMS Guideline, and the International Conference on Harmonization of Good Clinical Practice; our IRB adheres to the International Guideline for Human Research Safety (ICH-GCP). We obtained written signed consent from all patients or parents/legal guardians.

## Author’s contributions

HTA and DSA were employed of patients and collected the blood samples. The measurements of serum biomarkers were performed by HTA and HAH. The statistical analysis was carried out by MM. AA and MM wrote and edited the manuscript. All authors contributed to reading and approving the final version.

